# “The idea is to help people achieve greater success and liberty”: a qualitative study of expanded methadone take-home access in opioid use disorder treatment

**DOI:** 10.1101/2021.08.20.21262382

**Authors:** Leslie W. Suen, Stacy Castellanos, Neena Joshi, Shannon Satterwhite, Kelly R. Knight

**Author notes:** **Corresponding author:** Leslie Suen, MD, MAS, National Clinician Scholars Program, Philip R. Lee Institute of Health Policy Studies, University of California, San Francisco, 490 Illinois Street, Suite 7227, Box 0936, San Francisco, California 94158. **Declaration of interests:** None. **Author Contributions:** KRK and LWS conceived the design of the study. LWS, NJ, SC, and KRK conducted qualitative analyses. LWS took the lead in drafting the initial draft of the manuscript. All authors including LWS, NJ, SC, SS, and KRK contributed critical feedback to the writing of the manuscript. All authors approve the manuscript for publication.

## Abstract

**Background:** Prior to the COVID-19 pandemic, the United States (US) was already facing an epidemic of opioid overdose deaths. Overdose deaths continued to surge during the pandemic. To limit COVID-19 spread and to avoid disruptions in access to medications for opioid use disorder (MOUD), including buprenorphine and methadone, US federal and state agencies granted unprecedented exemptions to existing MOUD guidelines for Opioid Treatment Programs (OTPs), including loosening criteria for unsupervised take-home doses. We conducted a qualitative study to evaluate the impact of these policy changes on MOUD treatment experiences for providers and patients at an OTP in California.

**Methods:** We interviewed 10 providers and 20 patients receiving MOUD. We transcribed, coded, and analyzed all interviews to identify emergent themes.

**Results:** Providers discussed clinical decision-making processes and experiences providing take-homes. Implementation of expanded take-home policies was cautious. Providers reported making individualized decisions, using patient factors to decide if benefits outweighed risks of overdose and misuse. Decision-making factors included patient drug use, overdose risk, housing status, and vulnerability to COVID-19. New patient groups started receiving take-homes and providers noted few adverse events. Patients who received take-homes reported increased autonomy and treatment flexibility, which in turn increased likelihood of treatment stabilization and engagement. Patients who remained ineligible for take-homes, usually due to ongoing non-prescribed opioid or benzodiazepine use, desired greater transparency and shared decision-making.

**Conclusion:** Federal exemptions in response to COVID-19 led to the unprecedented expansion of access to MOUD take-homes within OTPs. Providers and patients perceived benefits to expanding access to take-homes and experienced few adverse outcomes, suggesting expanded take-home policies should remain post-COVID-19. Future studies should explore whether these findings are generalizable to other OTPs and assess larger samples to quantify patient-level outcomes resulting from expanded take-home policies.

## Introduction

The United States (US) was facing a deadly opioid overdose crisis when the COVID-19 pandemic began. Overdose deaths reached a record high of 75,000 in the twelve months preceding March 2020, and have continued surging during COVID-19.^1,2^ Buprenorphine and methadone are the two medications for OUD (MOUD) options proven to decrease mortality and increase care retention.^3,4^ Unlike buprenorphine, which can be accessed in office-based settings, methadone for OUD can only be dispensed through licensed Opioid Treatment Programs (OTPs) regulated by federal and state governments. Individuals are required to attend daily OTP visits, often with one unsupervised “take-home” dose per week when the OTP is closed. Prior to COVID-19, patients could receive privileges for unsupervised “take-home” doses after meeting strict guidelines posed by federal (e.g., Substance Abuse and Mental Health Services Administration (SAMHSA) and Drug Enforcement Agency (DEA)), and state agencies (e.g., the California Department of Health Care Services (DHCS)) (Table 1). Individuals meeting criteria could receive one additional take-home dose every 90 days of continuous treatment, and patients could receive up to 14 and 28 take-home doses after one and two years of treatment, respectively. However, due to the relapsing-remitting course of OUD, receiving higher amounts of take-homes for patients often took much longer.

**Table 1.** Overview of Federal and California State Guidelines Related to Methadone and Buprenorphine Take-home Prescriptions within Opioid Treatment Programs.

|  | **Federal criteria for take-home prescriptions from SAMHSA** | **State criteria for take-home prescriptions from DHCS** |
| --- | --- | --- |
| **Methadone** | All take-home decisions are determined by the medical director. Patients can receive an increase in take-home doses every 90 days in treatment up to a maximum of 2-week supply after one year of continuous treatment. After two years of continuous treatment, patients can receive up to 28-day supply of methadone if they meet criteria. The medical director uses the following criteria to determine permitting take-home privileges:<br>1. Absence of recent use of illicit or non-prescribed opioids, stimulants, benzodiazepines, alcohol, and other substances<br>2. Regular clinic attendance<br>3. Absence of serious behavioral issues at clinic<br>4. Absence of known recent criminal activity<br>5. Stability of home environment and social relationships<br>6. Sufficient length of time in treatment<br>7. Ability to safely store medications in a home environment<br>8. The rehabilitative benefit of decreasing frequency of clinic attendance outweighs potential risk of diversion | Patients must undergo regular medication monitoring with a prescribing clinician including randomly administered urine toxicology testing at least 8-12 times per year and attending monthly visits with a treatment counselor. |
| **Buprenorphine** | Recommended that patients starting buprenorphine have at least once weekly visits for the first month of treatment. The medical director can also use federal take-home requirements for methadone as a guideline for buprenorphine, though the time in treatment criteria does not apply. Patients can receive up to 28-day supply buprenorphine at a time. | Identical to Federal |
| **All Treatment** | Patients must undergo regular medication monitoring with a prescribing clinician including randomly administered urine toxicology testing at least 8-12 times per year and attending monthly visits with a treatment counselor. | Identical to Federal |

SAMHSA = Substance Abuse and Mental Health Services Administration
DHCS = California Department of Health Care Services

When the pandemic reached the US in early 2020, public health officials identified the congregate settings of OTPs as high-risk for COVID-19 spread. In March 2020, in efforts to reduce in-clinic crowding without disrupting MOUD access, SAMHSA provided federal guidance permitting blanket exceptions in all states, allowing up to 14 days of methadone take-homes for “unstable” patients and 28 days of take-homes for “stable” patients. OTPs had discretion in defining patients as “stable” or “unstable,” and these exemptions also provided other waivers related to urine toxicology monitoring and counseling.^5,6^ Notably, experts had advocated for expanded access to methadone take-homes for years to reduce barriers to care and to mitigate overdose deaths.^7,8^ These US regulatory changes to take-home guidelines represent an opportunity to evaluate expanded take-home regulations, ^9,10^ especially as similar lower-threshold methadone policies have seen success in other countries.^11–13^

We sought to qualitatively describe the MOUD treatment experiences of patients and providers at an OTP in San Francisco, California to inform future research and policy recommendations as the US shifts toward recovery from COVID-19.

## Methods

### Participants

We recruited and interviewed ten providers (including physicians, nurse practitioners, nurses, and behavioral health counselors) and 20 patients with OUD from one OTP in San Francisco. We used purposive sampling to interview providers and patients with diverse perspectives. We recruited providers via email and announcements made at weekly staff meetings, and we sampled to ensure a diversity of job disciplines. We asked providers to refer eligible patients to the study, and we aimed to recruit patients with diverse MOUD treatment experiences, duration, and treatment stability, and varied in COVID-19 exposure and risk. We oversampled patients who were new to MOUD treatment to increase likelihood of exposure to COVID-19-related regulatory changes.

### Data collection and analysis

We conducted interviews using a semi-structured guide focusing on MOUD access, utilization, and treatment experiences before and after COVID-19-related changes; impact of care practices and regulatory changes on drug use patterns, physical and mental health; and recommendations for care practice and regularity changes to maintain long-term. Interviews lasted ∼20-40 minutes. All participants provided verbal consent and received a $50 gift card for completing the interview. All provider participants completed interviews over Zoom videoconferencing software from August to September 2020. Patient participants completed the interview from September to November 2020, either over the phone or in person in an outdoor confidential space.

We audio-recorded, transcribed, and coded all interviews using Dedoose software. The analysis used a thematic approach with simultaneous data collection and analysis based on modified grounded theory methodologies.^14^ SC and KRK reviewed interview transcripts and analytic memos to develop a preliminary codebook. NJ coded transcripts and iteratively discussed with study investigators to add or revise existing codes. Once all interviews were coded, KRK and LWS independently reviewed interview transcripts and coding. They met regularly to discuss emergent themes until they reached consensus. The University of California, San Francisco institutional review board approved this study.

## Results

### Patient Populations Newly Eligible for Take-homes under Expanded Federal Exemptions

We interviewed ten OTP providers and 20 patients (Table 2). When SAMHSA released guidelines on expanded take-home restrictions, providers understood these policies were meant to reduce potential COVID-19 exposures by reducing in-person contact while maintaining MOUD access. These guidelines did not indicate specific criteria defining clinical stability or instability, introducing flexibility for individual OTPs to meet the need of their local areas during uncertain times.

**Table 2.** Demographics of Patients Receiving Medication for Opioid Use Disorder Participating in Individual Interviews (n=19)^1^

| Characteristic | N (%) or Median(IQR) |
| --- | --- |
| **Age, years** | 51 (41-60) |
| **Gender** |  |
| Man | 10 (53%) |
| Woman | 9 (47%) |
| Another gender identity | 0 (0%) |
| **Race/Ethnicity** |  |
| Black/African American | 9 (47%) |
| Hispanic/Latinx | 5 (26%) |
| Native American/American Indian | 2 (11%) |
| White | 5 (26%) |
| **Education** |  |
| Grammar school | 2 (11%) |
| High school degree or equivalent | 8 (42%) |
| Some college | 6 (32%) |
| Associate or bachelor's degree | 2 (10%) |
| Unknown | 1 (5%) |
| **Housing status in past 6 months** |  |
| In a permanent situation | 13 (68%) |
| In a vehicle parked on the street | 3 (16%) |
| In a shelter | 3 (16%) |
| In a temporary hotel | 2 (11%) |
| Stayed temporarily with friends/relatives | 3 (16%) |
| On the street or in a park | 5 (26%) |
| **Median Length in Opioid Treatment Program, months** | 36 (12-96) |
| **Prior treatment at Opioid Treatment Program** | 8 (42%) |
| **Current Medication for Opioid Use Disorder** |  |
| Buprenorphine | 2 (11%) |
| Methadone | 17 (89%) |
| Other | 0 (0%) |
| **Eligible for take-homes prior to COVID-19** |  |
| Yes, already receiving take-homes before COVID-19 | 2 (11%) |
| Yes, though not receiving take-homes before COVID-19 | 1 (5%) |
| No, started take-homes after COVID-19 | 11 (58%) |
| No, has not started take-homes after COVID-19 | 5 (26%) |

<sup>1</sup>Demographics not collected for one patient participant.

For this OTP, providers interpreted guidelines in ways that expanded take-homes for the following groups: 1) patients exposed to COVID-19, under investigation for COVID-19 symptoms, or COVID-19 positive needing to quarantine regardless of their drug use; 2) patients considered high risk for complications if they contracted COVID-19 (e.g., older adults, those with high risk or multiple comorbidities); 3) patients with well-controlled OUD and active stimulant use; and 4) patients who were stable in their MOUD treatment and newly housed in SIP hotels allowing safe storage for methadone. Providers highlighted that for patients with well-controlled OUD and active stimulant use who were previously ineligible for take-homes prior to COVID-19, the potential for take-home receipt represented a significant shift.

> *“The specific patient population that was affected the most by [changes in take-home policy] is people whose opioid use disorder is completely controlled and has been for years and they’re housed but they have a concomitant other substance use disorder, [which] specifically most commonly around here is either cocaine use disorder or methamphetamine use disorder. So [for] stimulant use disordered individuals who have active [stimulant] use, it would not be possible to give them take-homes [prior to COVID-19]*.*” [Provider A]*

### Providers were Cautious About Expanding Take-Homes for Patients

In addition to expanding to new groups, federal exemptions also provided individual OTPs discretion in deciding how many take-homes to provide eligible patients (within the boundaries of 14 and 28 days for unstable and stable patients, respectively) and how quickly to increase doses. Providers described being cautious and gradually increased take-homes for patients.

> *“We’re not just like blanketly giving tons of take-homes because we didn’t just suddenly say, ‘We’re giving them two weeks of take-homes*.*’ We’re doing things where we’re having them come in every other day, so we’re being quite judicious about it and making sure that we’re not giving people everything*.*” [Provider C]*

Reasons for being cautious included avoiding treatment destabilization and preventing misuse. A general culture of conservative dosing in OTPs and hearing about other OTP experiences of difficult outcomes from rapidly expanding take-home access informed providers to move slowly and cautiously. Providers described their clinical decision-making process, first by identifying newly eligible patients using a data-driven, experiential process. They discussed patient characteristics in teams and weighed the risks and benefits, based on patients’ drug use history, medical clinical comorbidities, vulnerability to COVID-19, and housing status. Providers then assessed the impact of take-homes to see if any adverse events occurred, and identified additional similar patients on a trial basis, incrementally increasing take-homes as opposed to systematic increases across the clinic population.

> *“One of the things we do is we talk about it as a case, so it’s not like an individual [provider] decision. Like I might go, ‘Hey, I think this person can handle take-homes, this is what they’re telling, they’re telling me they’re not using [opioids]*.*’ […] But the counselor said, ‘I think this person shouldn’t get take-homes. […] They just told me they were using benzos and heroin last week*.*’ I’m like, ‘Oh, it’s a completely different story than what [the patient] is telling me*.*’” [Provider C]*
>
> *“[Another provider said] ‘Well, here’s one client that I think will benefit, do you have any more?’ and based on his criteria of how, on that client, since I know my clients and their history now, I was like, ‘Okay, well, this person has the same story so let’s try them, this person has the same*.*’” [Provider D]*

Provider decisions around initiating or increasing take-homes centered on the balance of COVID-19 exposure risk and overdose risk. Providers referred to “getting to safety” when they could accumulate enough potential benefits of providing take-homes to justify the risks for patients. Providers struggled with finding the right balance of this risk assessment, as patients at highest risk for COVID-19 complications with multiple comorbidities were often also those at highest risk for opioid overdose.

> *“I think with COVID we’re trying to weigh the risk of coming in [to the clinic] for [high risk patients][…] And so having to come in every day versus once a week, dramatically increases their risk [of COVID-19 infection] if they’re coming to pick up their methadone everyday. But also someone who’s got COPD it’s at higher risk of overdose, especially if they’re handling their take-homes. So, how do you thread that needle where you don’t increase someone’s risk by giving them take-homes. So, we start off with a few, see if they can handle them and [then we can] increase*.*” [Provider C]*

Providers described relative contraindications to take-homes including concurrent, non-prescribed opioid, benzodiazepine, or significant alcohol use, or history of unsuccessful trials of receiving take-homes in the past. For these patients, providers considered the risks of take-homes to be higher than the risks of coming into clinic every day.

> *“I don’t really bring it up [take homes to the clinic team] if I know if they’re drinking [alcohol] very often, […] or if they’ve consistent benzo[diazepine] use and opiate use*.*” [Provider D]*

### Providers Highlighted Equity as a Guiding Principle for Deciding Take-Homes

Providers articulated a desire to be equitable and provide opportunities for take-homes.

> *“If I had a client and she was like ‘Oh, I want take-homes,’ and I’m like, ‘Okay, well, you have a pretty extensive history of continued [opioid] use so why don’t we give you a random UA and then we’ll take it from there*.*’ So those are more of the cases, is when they request it themselves*.*” [Provider D]*

An awareness about longstanding history of inequitable access to MOUD treatment especially among racial and ethnic minorities compelled providers to view their actions through an equity lens to ensure provision of take-homes was as fair and equitable.

> *“[In] a bigger system sort of perspective…[buprenorphine] is more accessible to white folk than Black and brown folk. And we don’t find that that is an acceptable state of being and we think that’s wrong, and we actively work to make sure that in our clinic that’s not the case*.*” [Provider A]*

### Patients and providers experienced increased patient autonomy, transparency, and treatment stabilization after receiving take-homes

Providers appreciated the flexibility they were given to implement the take-home policies.

> *“[We] don’t want to tie the hands of the counselor and [we] don’t want to make it so that the patients who have the loudest counselor get the most [take homes] – I don’t want that situation either. We want people to be able to say ‘Hey, this guy he really needs to be staying home right now. What can we do for him?’ […] The nice thing about the exceptions to the regulations that we have seen is that it allows us the flexibility to be creative in our treatment planning*.*” [Provider B]*

Both patients and providers noted how take-homes increased autonomy and flexibility for some, allowing them to attend to aspects of their lives like jobs, school, and caregiving, without jeopardizing their safety.

> *“The ability to use some real judgment, some human judgment—we’ve given some people take-home medications that has allowed them, I think, the same flexibility in their life. […] Like they’re doing other things with their time now. They’re seeing family or they’ve chosen to enter some kind of like quarantine pod, and they’re doing social things because we were able to sort of fudge it and say like, ‘We know you don’t meet [pre-COVID-19] criteria but that’s not the current criteria*.*’ […] I would say we’ve seen some positive changes*.*” [Provider E]*
>
> *“Well, it’s helped me a lot especially since I have a job, so I don’t have to worry about fitting going to the clinic into my daily schedule as well as my job and anything else that I do on a regular basis, like go to the gym and stuff*.*” [Patient A]*

For some providers, they noticed this flexibility in treatment plans led to increased transparency and honesty for patients.

> *“I feel like it’s proven in a lot of case[s] that [patients] could handle the methadone and it’s been nice to give them more trust in managing their methadone and not having to come in every day. I think it’s beneficial for the relationship*.*” [Provider F]*

Some patients reported that take-homes helped them to reduce their opioid use and stabilize in treatment, especially as getting to clinic daily was made harder with reduced public transportation options during COVID-19.

> *“[Having take-homes is] a blessing because I wasn’t [off heroin] before the COVID started. I wasn’t getting to the clinic every day; now I’m able to get there so it’s helped a lot to get stable, stabilized and get off of the drugs. […] Because I was trying not to do [heroin] before the COVID, and it was not that easy to stop even though I kept going up on my dose. […] But then when I started getting the take-homes and stuff, it made a big difference of drinking [methadone] every day. So then I didn’t have no more urges or anything. […] Not going in as often it was—you could tell the difference. So I’m grateful just to go in the few days that I go in and get take-homes. It’s working*.*” [Patient B]*

Patients remarked take-homes also allowed patients new autonomy in deciding when, where, and how much of their methadone to take. A patient receiving once-daily methadone dosing and waking up every morning in mild opioid withdrawal could now split their methadone dose to twice a day to ensure more even plasma concentrations and avoid withdrawal. Or a patient who experiences sedation immediately after taking methadone could opt to take their dose in the evening instead of in the morning, allowing them to pursue other activities during the day.

> *“[Having take-homes made] a lot of difference because I could take it any time, you know what I’m saying, when I need it*.*” [Patient C]*

### Patient Perceptions around Drug Exceptionalism and Lack of Transparency in Take-Home Policies

For patients with non-prescribed opioid, benzodiazepine, or heavy alcohol use who were still excluded from take-homes, they perceived continued lack of access to take-homes as inequitable, especially if providers couldn’t explain differential treatment due to concerns of violating patient privacy. Though for some patients, this lack of shared decision-making and transparency was interpreted as unfair allocation of take-homes. They perceived take-homes as a form of harm reduction, understood the risks of their drug use, and thought take-homes should be used as an incentive to remain engaged in treatment regardless of ongoing other drug use.

> *“I don’t understand what the give and take is about giving [take-homes]. I’m still coming every day; I’m still getting the same results [of not getting take-homes]. What’s the difference if I have something else in my system? [Getting take-homes] is harm reduction. What is the difference?” [Patient E]*

Even when patients with ongoing opioid, benzodiazepine, or heavy alcohol use understood the reasons behind why they couldn’t get take-homes, they still expressed frustration in response to what they perceived as differential treatment.

> *“It sucks because the only thing that’s prevented me ever from take-homes is benzos, that’s it. I don’t do any other drugs. I do methadone and I do Xanax. […] [W]hat’s frustrating is [my provider] tells me, ‘Look, if it was speed or cocaine or crack or anything else, we would be fine with giving you take-homes*.*’ But benzos, as you, I’m sure you know, ‘benzos’ is a very scary word to the medical field combined with any opiates or synthetic opiates. You have respiratory failure and major organ failure so they don’t like writing benzos with any kind of opiates; it’s a deadly cocktail*.*” [Patient F]*

### Considerations of What Policy Changes Should Be Maintained Post-COVID-19

Providers did not report observing increases in adverse events among patients who stayed in treatment and suggested that the expansion of take-home policies, especially regarding stimulant use, should continue after COVID-19 with careful monitoring. Further, providers were pleased with their method of expanding take-homes, especially as they had not subjectively seen many adverse outcomes. They preferred keeping flexibility for providers and individual OTPs in take-home decision-making for patients over prior policies:

> *“I haven’t seen any negativity or repercussions for the ones that we’ve selected. There are people that have been wanting take-homes even before COVID that are now pushing it even more and we still are not able to give it to them because of their using choices*.*” [Provider D]*

As more time passed during COVID-19, providers became more comfortable with balancing the risks and benefits of offering take-homes for patients, and these benefits warranted maintaining provider flexibility in take-home determinations after the COVID-19 pandemic.

> *“Keeping it as loose as possible so that individual clinics could do what they think is clinically appropriate feels like it would be safer than the old ways of doing things. […] The idea [is] to help people achieve greater success and greater liberty from us. And it’s been okay. We haven’t had any terrible stuff from that*.*” [Provider A]*

## Discussion

In this qualitative study of one OTP clinic’s experiences implementing take-home changes during COVID-19, expanded access to take-homes allowed providers new flexibility in their clinical decision making, which they described as experiential, team-based, and “judicious.” Providers balanced risks and benefits to “get to safety,” weighing individual factors such as drug use patterns, overdose risk, housing status, and vulnerability to COVID-19. Patients described increased treatment autonomy and flexibility with increased take-home receipt, including convenience, reduced burden of travel and risk for COVID-19 infection, and increased personal stabilization as benefits to take-home receipt. Some patients viewed their ongoing ineligibility for take-homes as a form of drug use exceptionalism and bias, and the perceived lack of transparency and shared decision-making increased dissatisfaction.

Large gaps in treatment access and capacity exist in the US, as <40% of patients with OUD access medication treatment.^15,16^ Other countries have expanded access to methadone by making it widely available through primary care and/or community pharmacy settings, in response to their own opioid overdose crises. One Canadian study in patients using injection drugs, low-barrier methadone for OUD was associated with lower all-cause mortality,^13^ while studies from Scotland and Australia reported substantially increased treatment capacity after methadone expansion to primary care/community pharmacy settings.^12,17,18^ Within the US, experts have called for similar expansion of methadone treatment outside of OTPs to increase access to methadone.^7,19^ Our findings suggest that maintaining guidelines that allow for expanded access to methadone takes-homes could increase access for patients who otherwise find it difficult to attend OTPs daily.

Patients and providers found take-homes increased treatment autonomy. Not having to attend methadone clinic daily meant freeing up energy and resources to tend to other responsibilities, including those related to employment, education, caregiving, and their other health conditions. Patients also had new freedoms in deciding when and how much of their methadone they would take to suit their physiologic needs and daily lives. Increasing patient treatment autonomy can enhance treatment engagement by increasing satisfaction, motivation, and self-empowerment.^20,21^

COVID-19 unfolded differently across US cities, creating natural experiments in how OTPs varied in their interpretation of whom they considered “unstable” or “stable” and how they applied expanded take-home policies accordingly. In our study, located in San Francisco, the city with the lowest death rate from COVID-19 of any US major city,^22^ providers described using a gradual, team-based approach to expanding take-homes. In six out of eight OTPs in the New England area defined criteria for take-home expansion narrowly and continued daily dosing for all new patients or those with positive drugs screens in the past 30 days.^23^ Meanwhile, in “hot spot” cities hit hardest by COVID-19 such as Seattle and the Bronx in New York City, OTPs described rapidly expanding take-homes to the majority of patients to reduce COVID-19 spread.^24,25^ Despite differences in approaches, the experiences of providers in our study mirror those described by providers in the Bronx, where clinicians sought to balance access to MOUD, risk of COVID-19 exposure, and risk of MOUD misuse and overdose, and used a team-based approach to balance safety and treatment equity.^24^

Our findings of increased benefits from take-home expansion without many apparent adverse events align with early reports from domestic and international methadone programs,^26–28^ and how flexibility in take-home exemptions allowed individual OTPs to decide which approaches best suited the needs of their patients, staff, and local environments.^29^ Our study is one of the first to describe the expansion of take-home privileges for patients with well-controlled OUD and active stimulant use, as this was one of the populations providers in this OTP defined as “stable” and therefore newly eligible for take-homes. OTPs traditionally are abstinence-oriented programs, previously disqualifying patients with any other non-prescribed drug or heavy alcohol use from take-homes even in the setting of well-controlled OUD.^30^ Prior studies suggested stimulant use during methadone treatment can lead to reduced pharmacologic efficacy of methadone, treatment retention, and abstinence from opioid use.^31,32^ However, these policies fail to acknowledge reasons why patients may use stimulants, including to combat the sedating effects of methadone, treat opioid withdrawal, or to improve daily functioning.^33^ Providing take-homes for patients with well-controlled OUD and ongoing stimulant use may be a reasonable approach, and future work should explore whether this patient population can continue receiving benefits of take-homes without meaningful harm.

Providers described employing equity as a guiding principle in their take-home decisions, being mindful of the racialized history of inequitable access to substance use treatment modalities.^34^ To this day, buprenorphine remains largely inaccessible to many Black and low-income communities.^35–37^ Understanding these historical contexts, providers in this study emphasized the importance of equitable access to MOUD treatment options and tried to extend this philosophy to take-home distribution.

Nevertheless, patients perceived differential approaches of take-home provision based on a drug use as a form of potential bias. Patients with stimulant use were eligible for take-homes while those with ongoing non-prescribed opioid, benzodiazepine, or heavy alcohol use were still disqualified. This difference inadvertently created a new form of drug exceptionalism in the clinic’s take-home prescribing guidelines, where like cannabis, patients using stimulants are no longer penalized like those using sedatives due to safety concerns. This drug hierarchy of privileging some drugs over others may have unintended consequences including increased stigma.^38^ This finding highlights how with increased decision-making flexibility for providers, the potential for perceived provider bias can increase in unpredictable ways, and counter-balancing measures may be needed to ensure equitable treatment and access for all patients.^39^

This study has several limitations. Generalizability of these findings is unclear as we collected data only from a single clinic, though reports from other OTPs across the country describe similar findings.^23–26^ We also conducted interviews during a period of frequent changes in clinic policy, and the impact of changes in take-home prescribing may not have yet reached steady state at the time of interviews. Despite these limitations, our study is one of the first to qualitatively describe the impact of expanded take-home prescribing practices on MOUD treatment with considerations for future research and policy.

## Conclusion

Unprecedented expansion of methadone regulatory policies during COVID-19 has forced OTPs to re-evaluate their clinical decision making and factor COVID-19 exposure and vulnerability when balancing safety risks in take-home assessments. Both providers and patients appreciated the flexibility and autonomy of expanded policies, with few reports of adverse outcomes. Maintaining expanded take-home regulations may meaningfully reduce barriers to MOUD access to curb overdose deaths, while aiming for equitable and transparent clinical decision-making practices.

## Data Availability

The dataset is not publicly available.

